# The epidemiology of systematic review updates: a longitudinal study of updating of Cochrane reviews, 2003 to 2018

**DOI:** 10.1101/19014134

**Authors:** Hilda Bastian, Jenny Doust, Mike Clarke, Paul Glasziou

## Abstract

**Background:** The Cochrane Collaboration has been publishing systematic reviews in the *Cochrane Database of Systematic Reviews* (*CDSR*) since 1995, with the intention that these be updated periodically.

**Objectives:** To chart the long-term updating history of a cohort of Cochrane reviews and the impact on the number of included studies.

**Methods:** The status of a cohort of Cochrane reviews updated in 2003 was assessed at three time points: 2003, 2011, and 2018. We assessed their subject scope, compiled their publication history using PubMed and *CDSR*, and compared them to all Cochrane reviews available in 2002 and 2017/18.

**Results:** Of the 1,532 Cochrane reviews available in 2002, 11.3% were updated in 2003, with 16.6% not updated between 2003 and 2011. The reviews updated in 2003 were not markedly different to other reviews available in 2002, but more were retracted or declared stable by 2011 (13.3% versus 6.3%). The 2003 update led to a major change of the conclusions of 2.8% of updated reviews (n = 177). The cohort had a median time since publication of the first full version of the review of 18 years and a median of three updates by 2018 (range 1–11). The median time to update was three years (range 0–14 years). By the end of 2018, the median time since the last update was seven years (range 0–15). The median number of included studies rose from eight in the version of the review before the 2003 update, to 10 in that update and 14 in 2018 (range 0–347).

**Conclusions:** Most Cochrane reviews get updated, however they are becoming more out-of-date over time. Updates have resulted in an overall rise in the number of included studies, although they only rarely lead to major changes in conclusion.

## Background

Systematic reviews use explicit formal methods to identify and analyse studies on specific research questions, and synthesise the findings of these studies. (1) Reviews’ findings can become outdated if they are overtaken by new studies or data, missing studies or errors in the review or included studies are identified, or methodology improves in critical ways. In a 2007 paper, Shojania et al reported on the need for updates in 100 systematic reviews from 1995 to 2005. (2) They concluded that around half may have been out-of-date, with a signal for required updating within two years of the evidence search in 11% of reviews. The median period without a signal suggesting a need to update was 5.5 years (95% confidence interval (CI), 4.6 to 7.6 years).

The intention to keep reviews up-to-date was a cornerstone of the Cochrane Collaboration. Named for Archie Cochrane, this international network aimed to achieve a goal he articulated by filling the need for: “a critical summary, by specialty or subspecialty, adapted periodically, of all relevant randomized controlled trials”. (3) The Cochrane Collaboration began publishing systematic reviews of the effects of healthcare interventions based on clinical trials in the *Cochrane Database of Systematic Reviews* (*CDSR*) in 1995. (4) Its organisers wrote in an introductory brochure in the 1990s that evidence reviews “must be prepared systematically and they must be kept up-to-date to take account of new evidence”. (5) A Cochrane review, they wrote, “is updated and amended as new evidence becomes available and errors are identified”.

The original expectation was that reviews would be updated at least annually, but in 2000 it was agreed that this was unsustainable, (6) and the expected update interval was changed to every two years, unless a reason was given for a different schedule. However, this also proved to be unsustainable, and in 2016 the organisation moved towards updating by perceived need or priority, preparing a consensus document on updating systematic reviews. (8) With the publication of a revision of the *Cochrane Handbook for Systematic Reviews of Interventions* (*Cochrane Handbook*) in 2019, there is no longer any default interval at which Cochrane reviews are judged to be out of date, and the possibility of retracting reviews deemed outdated will cease. (9)

Cochrane reviews are the result of a unique large-scale and long-term effort to stay systematically up-to-date with health and social care evidence across a broad spectrum of topics. This collection of reviews also provides an opportunity to study the practice of systematic reviewing and growth of evidence across time.

We assembled a cohort of Cochrane reviews, comprising all the reviews flagged as updated in the *CDSR* in 2003, and assessed these across 15 years at three time points: 2003, 2011, and 2018. The aims of our study were to describe the cohort of updated reviews in relation to all other Cochrane reviews that were available at the end of 2002 and, in 2011 and around 2018. We also charted and followed up on the updating history of the 2003 update cohort. We assessed how often the 2003 updates led to a major change in reviews’ conclusions. In addition, we charted the growth in the number of included studies in the reviews over time.

## Methods

### Study aim 1: Describe the cohort of reviews in the CDSR updated in 2003 in relation to all Cochrane reviews available at the end of 2002, 2011, and around 2018

The cohort was established in 2003 by searching each of the four issues of the *CDSR* published that year for reviews flagged as updated. The issue(s) in which each review was flagged as updated was recorded.

#### (a) Subject scope

Systematic reviews in different subject areas might become outdated more quickly than others. (2) Cochrane reviews are produced and maintained by editorial groups called Cochrane Review Groups (CRGs). These cover one or more subject areas, and have their own editorial policies. Differences in editorial practice could also affect updating. The number of CRGs grew throughout the early years of the Collaboration, with some merging in later years.

In order to gauge how similar in subject and editorial scope the 2003 cohort reviews might be to Cochrane reviews overall, we compared the spread of reviews among CRGS. A list of CRGs in 2019 was collected from the Cochrane website. (10) The names of the CRGs in each year from 1995 to 2002 were collected using the Internet Archive, (11) from a combination of archived issues of *Cochrane News* (12) and the Cochrane website. (13) The original CRG names were retained, and also normalised to the current CRG name, and mergers of groups were noted.

The CRG base for each of the reviews in the cohort was collected, noting where CRGs which have since been merged into another CRG. We compared the number of CRGs represented in the cohort with the number of CRGs in 2002 and 2019.

To compare the subject mix of the 2003 cohort with current Cochrane review production and maintenance, we gathered the number of new and updated reviews by CRG from the online Cochrane Library, collecting data for both 2017 and 2018 to reduce the impact of annual fluctuations. As the search function does not enable separation of new and updated reviews, the totals of new and updated reviews from 2017 to 2018 were collected from the dashboards on Cochrane’s website. (14)

#### (b) Likelihood of being updated

In 2011, data was collected to enable comparison of updating history between the 2003 cohort and other Cochrane reviews. In the time since 2003, the *CDSR* had moved from publication on CD-ROM to online publication, and had a change of publisher. (4,15) It became apparent that many previous versions of Cochrane reviews were no longer published in the *CDSR* and so we used PubMed to establish a cohort of reviews that had been published in the *CDSR* by the end of 2002, supplemented by a search of *CDSR* for reviews still dated pre-2003. The number of reviews that had been published by the end of 2002 was available from another project, (16) but that did not provide identifiers for those reviews.

In 2011, the status of each review that had been available in 2002 was identified using both PubMed and the latest version in the *CDSR*. There were three possible status designations of Cochrane reviews in 2011 at that time: ongoing, designated stable (no further update required), or withdrawn. Which of these had been applied to each review was recorded. As some of the reviews were no longer included in the CDSR without any withdrawal notice, we assigned that as a fourth category. The *CDSR* is unusual in having a large number of records withdrawn that could mean either the publication is out of date or that it “contains a major error”, and the reason for withdrawal is not routinely stated. (7) Another journal that also withdraws out-of-date publications classifies these all as retracted. (17) Opinions about whether all *CDSR* withdrawals constitute retractions vary. (18–20) In this study, the categories withdrawn and no longer included without notice were classified together as retracted.

Both PubMed and *CDSR* were used to establish whether the reviews had been updated between 2003 and 2011. Dates of updates were defined as the date of publication of a new citation version of the review, and date of search or incorporation of new data if no new cited version was published. When a version of a review involved only a software update, we did not count this as an update.

To compare updating and status between the reviews available in 2002 which had, and had not, been updated in 2003, we analysed the rate of ongoing, stable, and retracted reviews. We also analysed the proportions of ongoing reviews that had not been updated since 2002, those which were updated in 2003 only, and those which had been updated at least once between 2004 and 2011.

### Study aim 2: Describe the updating history of the cohort, major changes in conclusions in 2003, and the growth of included studies over time

All reviews flagged as updated in 2003 were collected. To assess the changes to the reviews’ conclusions, one author (HB) assessed all of these updates, and another (JD) assessed those published as updates in two of the four issues of the *CDSR*. Both authors agreed on a final group of reviews that had a major change in their conclusions following the update.

The first follow-up of the 2003 cohort of reviews was done in 2011. Using the first issue of *CDSR* in that year (published in April), the number of years from the review’s last reported search for eligible studies to April 2011 was recorded. After publication of the fourth and final issue of the *CDSR* for 2011, as complete as possible an updating history of the 2003 cohort was assembled. For each review in the cohort, PubMed was searched for previous records of the review. The version of the review published in issue 4 of 2011 was reviewed for information about updates in the three parts of the review that were expected to report the review’s history: What’s New, history, and notes. (21) As practice in what constituted an update had changed since 2003, two types of events were recorded as updates: the year of publication of a new citation of a version of the review in PubMed, and the year reported in the review for an update search or incorporation of new data even if no new cited version was published.

In March 2019, we added the year for each update from 2012 to the end of 2018. This was again based on PubMed searches and the information recorded in the reviews’ What’s New, history, and notes sections. In addition, we added the year the review was first published and the most recent PubMed identifier for the review.

This final data collection for the 2003 cohort alone included an assessment of the status of these reviews at the end of 2018, using the same categories as previously. However, we added a category for 2018: republished after previous retraction. For reviews that were stable, retracted, or republished after previous retraction, we recorded the years of these events.

When we originally identified reviews as updated in 2003, we recorded the number of studies included in that update, as well as the number included in the prior issue of the review. For one review, the updated version of the review was the only one available for assessment. We subsequently recorded the number of included studies in the versions of each review at the end of 2011 and the end of 2018.

### Data management and analysis

One author (HB) undertook all data collection, curation, and visualisation. Data for the first collection in 2003 was originally recorded in document tables, and added to an Excel spreadsheet in 2019. All other collections were recorded in Excel. Data was analysed using RStudio 1.1463 running R 3.5.2. (9) Packages tidyverse and (24) reshape2 (25) were used for analyses and data visualisation. Summary statistics were used to describe the cohort. Data for this project, including analytic code, are available at Github. (26)

## Results

### 2003 cohort in perspective: subject scope

We identified 1,532 Cochrane reviews via PubMed and the *CDSR* that were available at the end of 2002. This was fewer than the number reported by the Cochrane Collaboration since the beginning of the *CDSR* in 1995 (1,558). (16) Some reviews had been indexed in PubMed after 2002 (n = 13), and this is likely to be the case for more of the shortfall of 26 reviews. Other reviews may have been retracted prior to the indexing of the *CDSR* in PubMed in 2000. (15) A total of 177 reviews were flagged as updated in the *CDSR* in 2003. As the first version of four of these was also published in 2003, the update rate in 2003 of 1,532 reviews indexed in PubMed to the end of 2002 was 11.3%.

There were 49 Cochrane Review Groups (CRGs) with editorial responsibility for Cochrane reviews in 2002, compared with 53 in 2019. Table 1 shows the rise in the number of CRGs between 1995 and 2002, as well as annual review production based on Cochrane-supplied data and the years of first publication of the 173 reviews in the update cohort reviews that had been published by the end of 2002. CRGs had a median of two reviews updated in 2003, ranging from none to 27 (interquartile range (IQR): 3).

**Table 1.** Annual number of CRGs and new Cochrane reviews, 1995–2002, with year of first publication for the 2003 update cohort.

| Year | Number of CRGs | Number of reviews |  | Year of first publication of the reviews in the 2003 update cohort |  |
| --- | --- | --- | --- | --- | --- |
|  |  | New | Cumulative total | Number | % of all reviews<sup>1</sup> |
| 1995 | 18 | 65 | 65 | 10 | 15.4 |
| 1996 | 22 | 105 | 170 | 13 | 7.7 |
| 1997 | 30 | 139 | 309 | 15 | 4.9 |
| 1998 | 45 | 208 | 517 | 21 | 4.1 |
| 1999 | 46 | 184 | 701 | 20 | 2.9 |
| 2000 | 48 | 264 | 965 | 37 | 3.8 |
| 2001 | 49 | 309 | 1274 | 36 | 2.8 |
| 2002 | 49 | 284 | 1558 | 21 | 1.3 |
| Total | *n.a.* | 1,558 | *n.a.* | 173<sup>2</sup> | *n.a.* |

<sup>1</sup> Percentage of total number of Cochrane reviews available in that year.
<sup>2</sup> This excludes four reviews in the 2003 update cohort that were also published for the first time in 2003.

Only four CRGs had been in existence for less than four years in 2002, and 19 were no more than five years old. At that time annual updating of Cochrane reviews was expected, but only 42 of the 49 CRGs (85.7%) flagged reviews as updated in 2003, and only 11.3% of the reviews published by the end of 2002 were updated during 2003.

To gauge how similar the subject scope of reviews in the 2003 update cohort are to the current subject scope of Cochrane reviews, we compared the spread of reviews among CRGs in the 2003 update cohort with the spread of new and updated reviews published in 2017/18 (Table 2). The *CDSR* does not enable breaking these down into new and updated reviews but the 2017/2018 totals reported in Cochrane dashboards online show that 44.8% of these reviews were updates. (14) (The dashboards report fewer total new and updated reviews for 2017/2018: 1,353 versus 1,369 returned by *CDSR* search).

**Table 2.** Proportion of reviews per CRG: 2003 update cohort and all new and updated reviews in 2017/18.

| **Individual CRGs** | **2003 update cohort**<br>**(n = 177)** | **All reviews**<br>**2017/2018**<br>**(n = 1,369)** |
| --- | --- | --- |
| Number of reviews per CRG (median) | 3 | 19 |
| Number of reviews per CRG (range) | 1 to 27 (IQR 3) | 2 to 95 (IQR 23) |
| Proportion of the CRG's reviews (median) | 1.7% | 1.4% |
| Proportion of the CRG's reviews (range) | 0.6% to 15.3% | 0.1% to 6.9% |

Cochrane reviews in the 2003 update cohort were distributed across a narrower range of CRGs, and therefore topic areas, than the new and updated reviews in 2017/18 (Table 2). The extent of this shift is illustrated in the case of the oldest CRGs. Of the 18 CRGs that had formed by 1995, two later merged. Those 17 CRGs were responsible for 57.6% of the 2003 update cohort (102 of 177 reviews), and 48.0% of the new and updated reviews in 2017/18 (657 of 1,369 reviews). The individual CRGs, based on their 2019 names and grouping, are shown with their proportion of reviews in both time periods in Figure 1.

**Figure 1.**
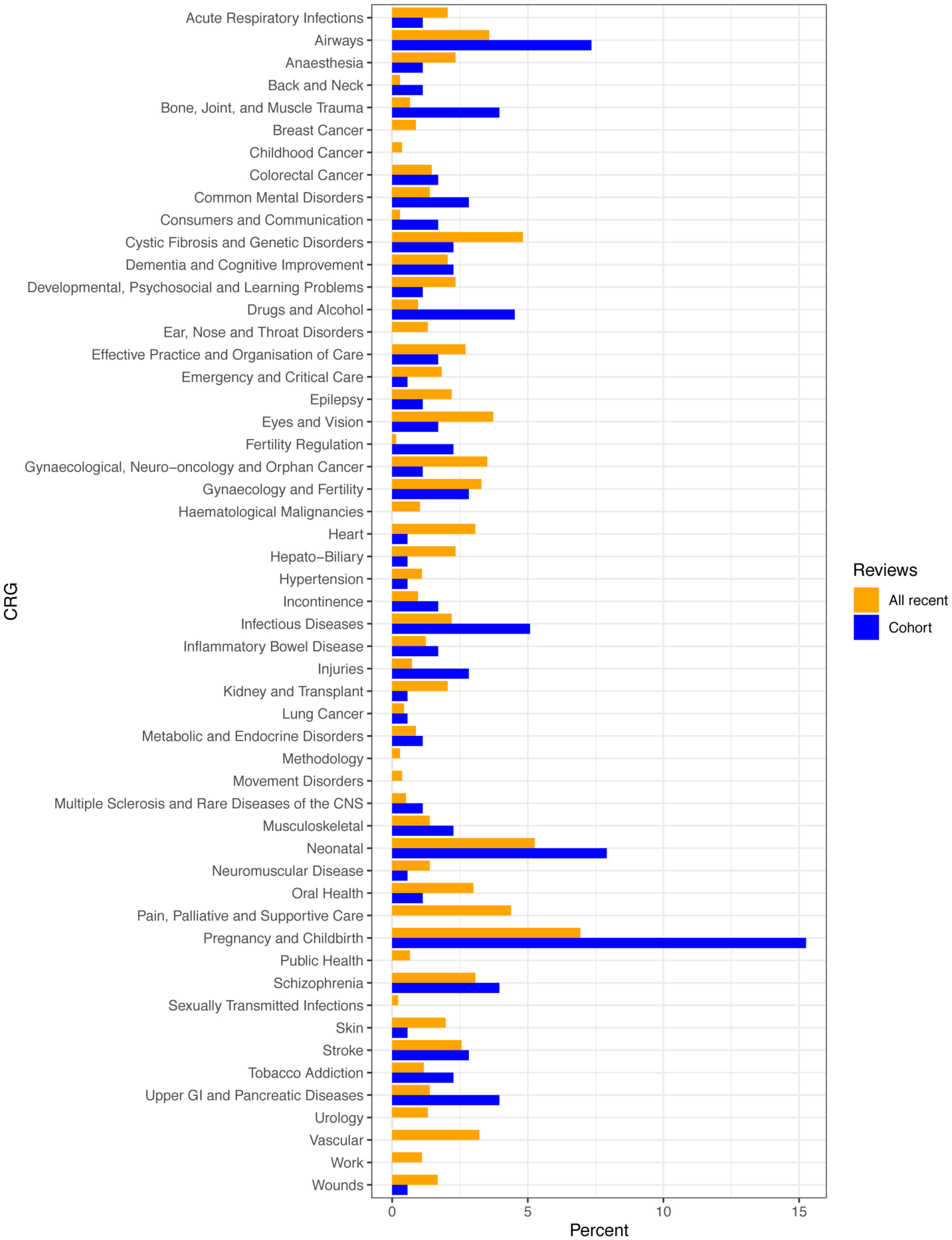
Proportion of reviews per CRG: 2003 update cohort and new and updated reviews in 2017/18. Note: Two individual CRGs from 2003 had merged by 2019 and are treated as merged in 2003 for this comparison.

### 2003 cohort in perspective: likelihood of being updated

The updating status of the 1,532 reviews published by the end of 2002 was categorised in 2011 to show whether each review was ongoing, stable (no longer being updated), or retracted at that time, and whether or not they had been updated at least once between 2003 and 2011. The retracted reviews included those with a withdrawal notice as well as six reviews that were no longer in the *CDSR*, with no record to explain their absence.

We compared the 173 reviews in the 2003 update cohort for which the original version had been published before 2003 with the other reviews that were available at the end of 2002 (Table 3).

**Table 3.** Comparison of 2002 reviews updated in 2003 (*n* = 173) with update status of other reviews available in 2002 (*n* = 1532).

| Review status by the end of 2011 | Updated in 2003 |  | No 2003 update |  | Total |  |
| --- | --- | --- | --- | --- | --- | --- |
|  | No. | % | No. | % | No. | % |
| Ongoing | 150 | 86.7 | 1,274 | 93.7 | 1,424 | 93.0 |
| Stable | 11 | 6.4 | 26 | 1.9 | 37 | 2.4 |
| Retracted | 12 | 6.9 | 59 | 4.3 | 71 | 4.6 |
| **Total reviews** | **173** | **100** | **1,359** | **100** | **1,532** | **100** |
| Ongoing with no update since 2002 | – | – | 237 | 18.6 | 237 | 16.6 |
| Ongoing and updated in 2003 only | 22 | 14.7 | – | – | 22 | 1.5 |
| Ongoing and updated at least once between 2004 and 2011 | 128 | 85.3 | 1,037 | 81.4 | 1,165 | 81.8 |
| **Total ongoing reviews** | **150** | **100** | **1,274** | **100** | **1,424** | **100** |

The proportion of 2002 Cochrane reviews that were ongoing in 2011 and had been updated at least once between 2004 and 2011 was high, whether they had been updated in 2003 (85.3%) or not (81.4%), suggesting that the 2003 update cohort is not markedly different to the other 2002 reviews in regard to subsequent updating over the following eight years. However, the 2003 cohort had higher rates of retraction and being declared stable than other reviews from 2002 by 2011 (13.3% versus 6.3%). By 2019, the overall rate of non-retracted Cochrane reviews being declared stable was 6.6% *[Chapter 4]*, compared with 6.8% for our 2003 update cohort (11 of 161 non-retracted reviews). In regard to the number of included studies, the 2003 update cohort include two reviews with no included studies in 2011 (1.1%), which was a lower proportion than Yaffe et al found for all Cochrane reviews in 2010 (8.7%). (27)

### Updating history of the 2003 update cohort from publication to 2018

The reviews in the 2003 cohort were first published as full Cochrane reviews between 1995 and 2003, with a median time since first publication of 18 years by the end of 2018 (range: 15 to 23 years; IQR 3) (Figure 2).

**Figure 2.**
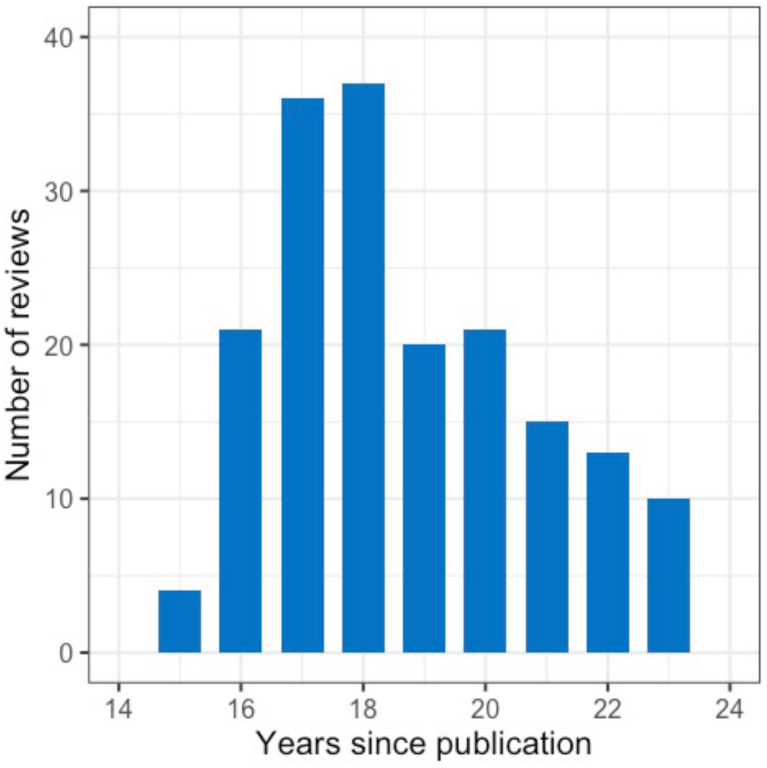
Years since first publication of reviews to the end of 2018 (n = 177).

From first publication until 2018, the median number of updates per review was three (range: 1 to 11, IQR 2) (Figure 3).

**Figure 3.**
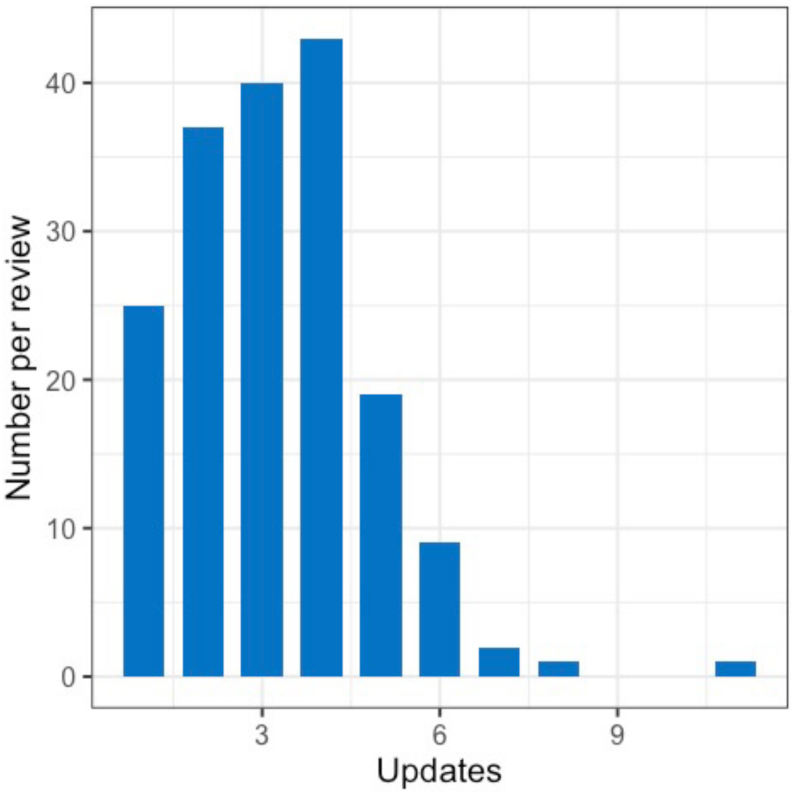
Number of updates per review to 2018 (n = 177).

Among the 177 reviews in the 2003 update cohort, 150 were ongoing in 2018: the other 27 were either retracted (n = 15) or designated stable (n = 12). One review had been retracted in 2010, but was updated and republished the following year, and is included among the 150 ongoing reviews.

The median time to each update of these reviews was three years (Table 4). The shortest was zero in the case of the first update, as some reviews were updated during the year they were published. The longest interval between updates was 14 years between the first and second updates of one review.

**Table 4.** Median time in years to first and subsequent updates for the 150 ongoing reviews from the 2003 update cohort.

| No.<br>(%) | Time to update in years – median (range) |  |  |  |  |  |  |  |  |  |  |
| --- | --- | --- | --- | --- | --- | --- | --- | --- | --- | --- | --- |
|  | 1<sup>st</sup> | 2<sup>nd</sup> | 3<sup>rd</sup> | 4<sup>th</sup> | 5<sup>th</sup> | 6<sup>th</sup> | 7<sup>th</sup> | 8<sup>th</sup> | 9<sup>th</sup> | 10<sup>th</sup> | 11<sup>th</sup> |
| 150<br>(100) | 3<br>(0-8) |  |  |  |  |  |  |  |  |  |  |
| 135<br>(90.0) |  | 3<br>(1-14) |  |  |  |  |  |  |  |  |  |
| 107<br>(71.3) |  |  | 3<br>(1-8) |  |  |  |  |  |  |  |  |
| 72<br>(48.0) |  |  |  | 3<br>(1-10) |  |  |  |  |  |  |  |
| 32<br>(21.3) |  |  |  |  | 3<br>(1-8) |  |  |  |  |  |  |
| 13<br>(8.7) |  |  |  |  |  | 3<br>(1-9) |  |  |  |  |  |
| 4<br>(2.7) |  |  |  |  |  |  | 3<br>(1-5) |  |  |  |  |
| 4<br>(2.7) |  |  |  |  |  |  |  | 3<br>(1-5) |  |  |  |
| 2<br>(1.3) |  |  |  |  |  |  |  |  | 2 |  |  |
| 1<br>(0.7) |  |  |  |  |  |  |  |  |  | 2 |  |
| 1<br>(0.7) |  |  |  |  |  |  |  |  |  |  | 2 |

Figure 4 charts the ongoing reviews in 2018 by the years since their last update. A total of 36 (24.0%) reviews had not been updated in the previous 10 years. Further analyses are included in Table 5, together with other analyses of the currency of the 2003 update cohort.

**Table 5.**
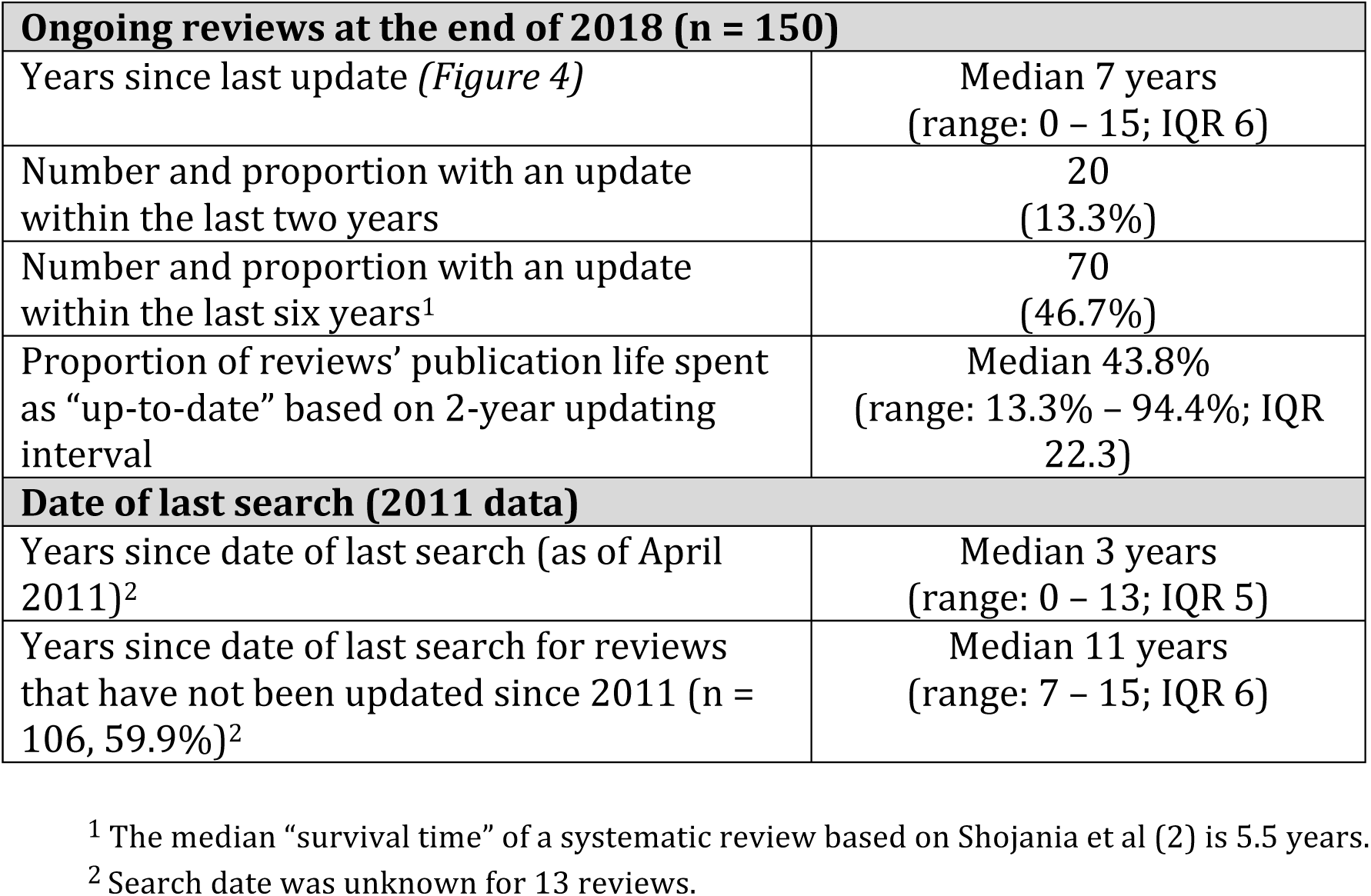
Additional indicators of review currency.

| **Ongoing reviews at the end of 2018 (n = 150)** |  |
| --- | --- |
| Years since last update (*Figure 4*) | Median 7 years<br>(range: 0 – 15; IQR 6) |
| Number and proportion with an update within the last two years | 20<br>(13.3%) |
| Number and proportion with an update within the last six years<sup>1</sup> | 70<br>(46.7%) |
| Proportion of reviews' publication life spent as "up-to-date" based on 2-year updating interval | Median 43.8%<br>(range: 13.3% – 94.4%; IQR 22.3) |
| **Date of last search (2011 data)** |  |
| Years since date of last search (as of April 2011)<sup>2</sup> | Median 3 years<br>(range: 0 – 13; IQR 5) |
| Years since date of last search for reviews that have not been updated since 2011 (n = 106, 59.9%)<sup>2</sup> | Median 11 years<br>(range: 7 – 15; IQR 6) |

<sup>1</sup> The median "survival time" of a systematic review based on Shojania et al (2) is 5.5 years.
<sup>2</sup> Search date was unknown for 13 reviews.

**Figure 4.**
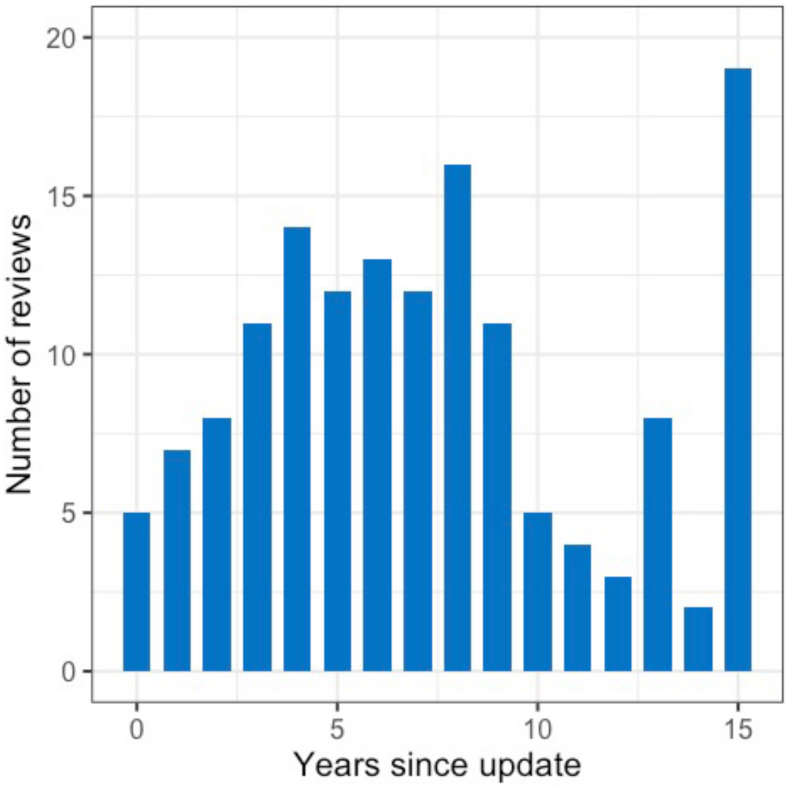
Ongoing reviews by years since last update, 2018 (n = 150).

### Impact of updating over time

In 2003, the updating of a review resulted in major changes to its conclusions for five reviews (2.8%). We compared the number of included studies in the updated review with the number in its prior version (excluding one review had been retracted without leaving a copy of the original review in *CDSR*, leading to a single missing baseline value). Some updates did not lead to the inclusion of further studies, but the number of included studies grew substantially over time (median: 8 from the version before the 2003 update to 14 in the version of the review at the end of 2018) (Figure 5).

**Figure 5.**
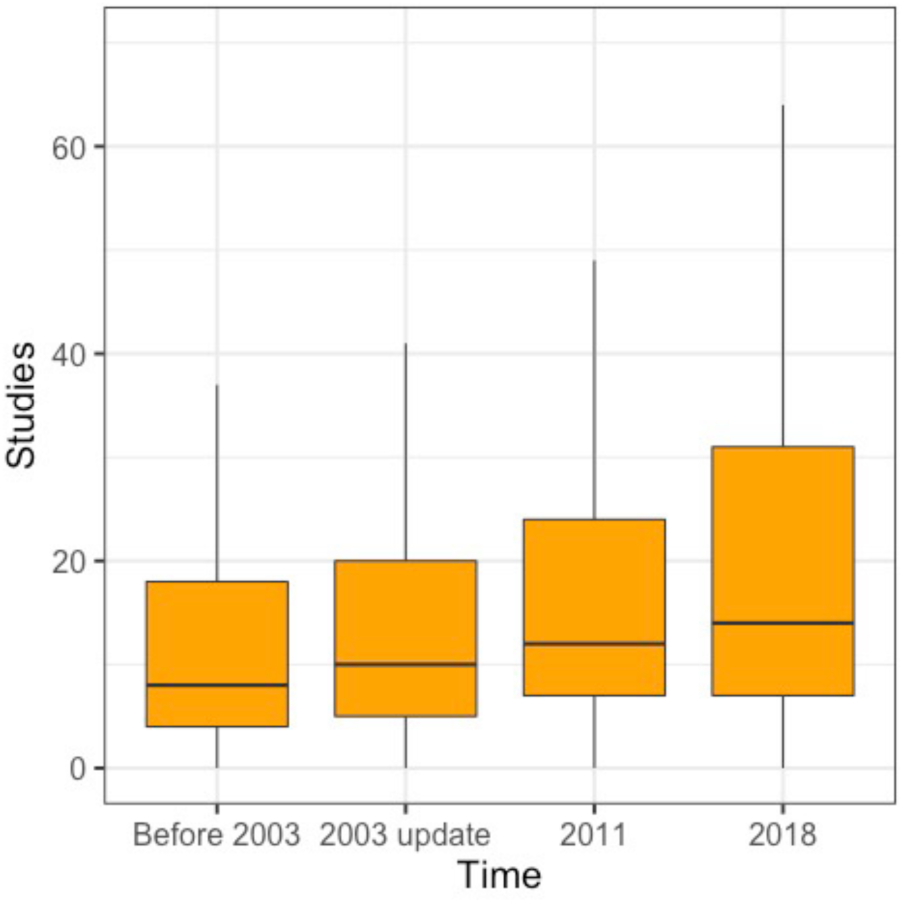
Included studies in reviews: baseline, 2003, 2011, and 2018. Notes: Outliers not displayed; 1 missing value at baseline (before 2003).

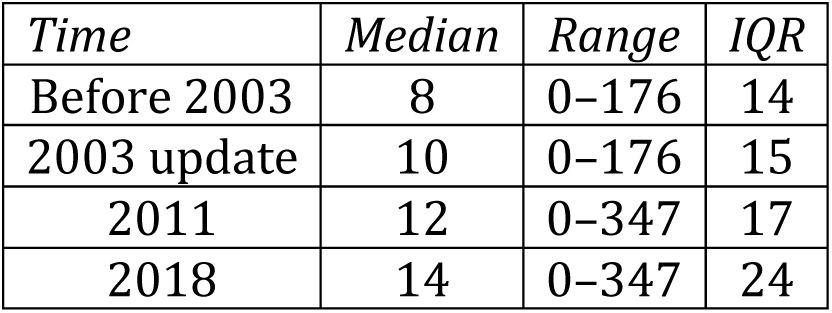

| *Time* | *Median* | *Range* | *IQR* |
| --- | --- | --- | --- |
| Before 2003 | 8 | 0–176 | 14 |
| 2003 update | 10 | 0–176 | 15 |
| 2011 | 12 | 0–347 | 17 |
| 2018 | 14 | 0–347 | 24 |

## Discussion

Our study describes the updating history over 15 years of a cohort of 177 Cochrane reviews that were first updated in 2003. For most of this period, the Cochrane updating policy was a recommended two-year interval until a review was regarded as out of date. By that measure, reviews in this cohort could be considered out of date for more than half of their publication life. However, in another study Shojania et al concluded that the median life of a systematic review before it became outsided was 5.5 years (CI, 4.6 to 7.6 years), based on a sample of 100 systematic reviews. As the median time between updates for the Cochrane reviews in our 2003 update cohort was three years, most of this cohort were likely to be up-to-date most of the time.

However, this cohort has been getting more out of date over time. The median time to first update was three years, but by the end of 2018, more than half the reviews were more than seven years since their last update (range 0–15 years). This is reflective of Cochrane reviews more generally, as more new reviews are published than updated and the legacy of existing reviews now exceeds 8,000. If indeed the interval between review updates continues to increase in the long term, or more and more reviews are never updated, up-to-date Cochrane reviews will become the exception, not the rule.

Until recently, Cochrane policy allowed for withdrawing a review from publication when it was seriously out of date. This resulted in a high retraction rate for our cohort (6.9%), relative to its fellow Cochrane reviews that were not updated in 2003 (4.3%). With the introduction of a new update classification system that aims to phase out the withdrawal of outdated reviews, (28) the retraction rate in future is likely to be considerably lower. The proportion of reviews showing as apparently current in the *CDSR* but which are seriously outdated will be correspondingly higher.

Our cohort had a higher rate of updates than other Cochrane reviews from the early 2000s. In addition, they included a particularly low rate of “empty” reviews in 2011 (1.1% without included studies) compared to the 8.7% rate Yaffe et al reported for all Cochrane reviews in 2010. (27) In our study, the median number of included studies prior to the 2003 update was eight. Mallett and Clarke reported that the median number of included studies in all Cochrane reviews at the beginning of 2001 was six. (29) The 2019 *Cochrane Handbook* advises authors of reviews to consider, among other things, whether new eligible studies are likely to be found before deciding to update. (9) Our results are consistent with Cochrane groups already concentrating updating effort on research questions that were generating new studies.

Our results for the earlier years are similar to the results from studies with shorter follow-up than ours. We identified 13 other studies of updating in Cochrane reviews or protocols that addressed some similar outcomes (overview in Appendix). (30– 42) Mean or median times to updates were comparable to those in our study. Our finding that a major change in conclusion after update is rare (2.8%) is also consistent with others’ results. Jaidee et al found a 2.0% rate of major changes in their conclusions at the first update of 101 Cochrane reviews. (39) Bashir et al found a 3.9% change in their conclusions in 8 out of 204 reviews with a meta-analysis for a primary outcome. (42) The highest rate was found by French et al, (34) who found a 9.1% change in review conclusions in 23 out of 254 reviews, but these changes were not necessarily major.

There have been two studies with similar analyses of updating non-Cochrane evidence syntheses. The National Institute for Health and Care Excellence (NICE) reported on updating of their systematic review-based clinical practice guidelines. (43) For 11 guidelines, the median time to update was 5.3 years (range: 3.3 to 6.5 years), with major changes in recommendations in six of them. Peterson et al studied 41 comparative effectiveness reviews of drugs, finding a median time to update of just over two years. (44)

Our study has several important strengths, particularly its long-term follow-up and open data that could enable others to extend this longitudinal study. The collection of cross-sections of data in 2003, 2011, and 2017/2018 was valuable. By maintaining a historical collection of data and using PubMed as well as the *CDSR*, we established that the updating and version history of Cochrane reviews in the *CDSR* has important gaps. While most older versions of reviews are online in the *CDSR*, some complete reviews and some update versions of reviews have been removed from the *CDSR* without leaving any public record other than PubMed. Some updated versions of Cochrane reviews were never submitted to PubMed. In some cases this may be in error, but it may also be a matter of policy. (7) This has left no complete public record of all Cochrane reviews, their updates, versions, and retractions. Not submitting updates for indexing at PubMed has critical implications for searches: reviews with misleadingly older dates will not be retrieved by searches that are limited to more recent records.

The inadequate historical record was one of the limitations of our study. We also relied on manual collection of data and quality assurance by a single author. In assessing the growth of included studies, we did not collect data on eligible studies awaiting inclusion and we do not know if this would have affected our results.

Our study raises some issues for Cochrane and others to consider. We could find the date of last search clearly reported in 92.7% of the reviews in 2011. In a study published in 2013, Beller et al found that only 90.0% of a sample of systematic reviews from PubMed reported the date of last search. (45) The availability of that date is critical for users to assess the currency of evidence and the adequacy of overlapping periods in update searches.

In 2002, Koch drew attention to the poor quality of update reporting in Cochrane reviews, (31) and we found this to be an ongoing problem. Contributing factors include missing versions of Cochrane reviews in the *CDSR*, and reviews not carrying forward events from “What’s New” and other notes sections into the history of each subsequent version of the review as Cochrane advises. (7) Automation of that process could improve this situation.

A further critical gap is the lack of reporting of searches undertaken before it is determined that an update is needed. Although searches that found no new studies were generally reported as updates to the reviews in early years when annual or two-yearly updates were expected, these are no longer as visible. (8,9) Transparency of this element of updating activity is important for users, including other systematic reviewers and producers of clinical practice guidelines and health information generally. A survey of health organisations producing systematic reviews (and often clinical practice guidelines) by Garritty et al in 2010 had 114 respondents (30% of them from Cochrane). Only eight of the organisations did not have any updating activity, but resources for updating for those that did were stretched, and the groups regarded too many of their reviews as too far out of date. Sharing the results of searches is critical in this context to prevent large numbers of groups using resources on the same futile searches.

In 2010, two of us advocated that major changes were needed to keep up with the evidence, given the massive rise in clinical trials and increasing complexity of systematic review methods, with constrained resources. (16) Prioritisation of systematic reviews, reduction of avoidable waste, and “leaner and more efficient methods of staying up-to-date” were stressed. Since then, the rise in clinical trials has escalated: ClinicalTrials.gov amassed over 96,000 trials in its first decade to 2010, and the total nearly doubled in the next five years. (46,47) It now stands at over 320,000 (as of November 2019).

Progress in streamlining methods and avoiding waste are not moving as fast. Methodological expectations for Cochrane reviews have increased, (9,48) and trial registry entries have been added to Cochrane’s trial database as the organization grapples with the implications of incorporating unpublished trials and data. (49) There has, however, been a concerted effort to develop and implement methods for prioritisation of updating for Cochrane reviews. (8) With this transition to targeted updating, new questions about processes and impact need to be tackled. Are the right systematic reviews being updated? Are people being harmed by reliance on outdated systematic reviews?

## Data Availability

Data and supporting files, including analytic code, are available at GitHub

https://github.com/hildabast/cochrane-updating/

## Author contributions

HB initiated the study, undertook the data collections in 2003, 2011, and 2018, curated, analysed and visualised data, and drafted the manuscript. HB and JD jointly conceptualised the establishment of the cohort in 2003 and agreed on reviews with major changes in conclusions. HB, JD, PCG, and MC participated in conceptualising the 2011 follow-up. The 2018 follow-up was initiated and conceptualised by HB. All authors contributed to interpretation of the data and critical revision of the manuscript.

## Acknowledgments

We are grateful to Robyn Brown, who participated in the first assessment of the cohort in 2003. We are also grateful to Claire Allen, who in 2009 provided the data underlying the chart of Cochrane reviews from 1995 to 2008 then published on the Cochrane website.

## Abbreviations

CDSR: *Cochrane Database of Systematic Reviews* (journal component of The Cochrane Library)
CI: Confidence interval
CRG: Cochrane review group
IQR: Interquartile range
PRISMA: Preferred Reporting Items for Systematic Reviews and Meta-Analyses

